# Integrating 4 Measures to Evaluate Physical Function in Patients with Cancer (In4M): Protocol for a prospective study

**DOI:** 10.1101/2023.03.08.23286924

**Authors:** Gita Thanarajasingam, Paul G. Kluetz, Vishal Bhatnagar, Abbie Brown, Elizabeth Cathcart-Rake, Matthew Diamond, Louis Faust, Mallorie H. Fiero, Scott F. Huntington, Molly Moore Jeffery, Lee Jones, Brie N. Noble, Jonas Paludo, Brad Powers, Joseph S. Ross, Jessica D. Ritchie, Kathryn J. Ruddy, Sarah E. Schellhorn, Michelle E. Tarver, Amylou C. Dueck, Cary P. Gross

**Affiliations:** Division of Hematology, Mayo Clinic, Rochester, Minnesota, USA; U.S. Food and Drug Administration, Silver Spring, Maryland, USA; Health Education and Content Services, Mayo Clinic, Rochester, Minnesota, USA; Department of Medical Oncology, Mayo Clinic, Rochester, Minnesota, USA; Division of Health Care Delivery Research, Kern Center for the Science of Health Care Delivery, Mayo Clinic, Rochester, Minnesota, USA; Department of Internal Medicine, Yale School of Medicine, New Haven, Connecticut, USA; Yale’s Cancer Outcomes, Public Policy, and Effectiveness Research (COPPER) Center, Yale School of Medicine, New Haven, Connecticut, USA; Department of Emergency Medicine, Mayo Clinic, Rochester, Minnesota, USA; Patient advocate, Arlington, Virginia, USA; Department of Quantitative Health Sciences, Mayo Clinic, Phoenix, Arizona, USA; CancerHacker Lab, Boston, Massachusetts, USA; Yale-New Haven Center for Outcomes Research and Evaluation (CORE), New Haven, Connecticut, USA; Department of Health Policy and Management, Yale School of Public Health, New Haven, Connecticut, USA; Department of Chronic Disease Epidemiology, Yale School of Public Health, New Haven, Connecticut, USA

## Abstract

**Introduction:** Accurate, patient-centered evaluation of physical function in patients with cancer can provide important information on the functional impacts experienced by patients both from the disease and its treatment. Increasingly, digital health technology is facilitating and providing new ways to measure symptoms and function. There is a need to characterize the longitudinal measurement characteristics of physical function assessments, including clinician-reported physical function (ClinRo), patient-reported physical function (PRO), performance outcome tests (PerfO) and wearable data, to inform regulatory and clinical decision-making in cancer clinical trials and oncology practice.

**Methods and analysis:** In this prospective study, we are enrolling 200 English- and/or Spanish-speaking patients with breast cancer or lymphoma seen at Mayo Clinic or Yale University who will receive standard of care intravenous cytotoxic chemotherapy. Physical function assessments will be obtained longitudinally using multiple assessment modalities. Participants will be followed for 9 months using a patient-centered health data aggregating platform that consolidates study questionnaires, electronic health record data, and activity and sleep data from a wearable sensor. Data analysis will focus on understanding variability, sensitivity, and meaningful changes across the included physical function assessments and evaluating their relationship to key clinical outcomes. Additionally, the feasibility of multi-modal physical function data collection in real-world patients with cancer will be assessed, as will patient impressions of the usability and acceptability of the wearable sensor, data aggregation platform, and PROs.

**Ethics and dissemination:** This study has received approval from IRBs at Mayo Clinic, Yale University, and the U.S. Food & Drug Administration. Results will be made available to participants, funders, the research community, and the public.

**Registration Details:** The trial registration number for this study is NCT05214144

**Strengths & Limitations:**

- This study addresses an important unmet need by characterizing the performance characteristics of multiple patient-centered physical function measures in patients with cancer
- Physical function is an important and undermeasured clinical outcome. Scientifically rigorous capture and measurement of physical function constitutes a key component of cancer treatment tolerability assessment both from a regulatory and clinical perspective.
- This study will include patients with lymphoma or breast cancer receiving a broad range of cytotoxic chemotherapy regimens. While recruitment will occur at two academic sites, patients who ultimately receive treatment at local community sites will be included.
- A patient-centered health data aggregating platform facilitates the delivery of patient-reported outcome measures and collection of wearable data to researchers, while reducing patient burden compared to traditional patient-generated data collection and aggregation methods
- Heterogeneity in patient willingness or comfort engaging with mobile products including smartphones and wearables, enrollment primarily at large academic centers, and the modest sample size are potential limitations to the external validity of the study

## INTRODUCTION

Cancer clinical trials have long emphasized important metrics of tumor response and survival rates to evaluate the benefit of cancer trials. However, there has been increasing recognition of the importance of systematically assessing how patients feel and function – tolerability – while on treatment.^1^ Disease-related symptoms, physical function, and toxicity (i.e. side effects from treatment) are core outcomes that have been identified by the United States Food & Drug Administration to inform the safety, tolerability and efficacy of an investigational cancer therapy^2 3^.

Physical function (PF) is defined as the ability to carry out day-to-day activities that require physical effort^4^. Symptoms related to a patient’s underlying cancer as well as treatment-related toxicity can impact PF. PF can be assessed using multiple complementary approaches. These include clinician-or investigator-reports (e.g. Eastern Cooperative Oncology Group [ECOG] performance status [PS]^5^), patient-reported outcome measures (PROs; e.g., questionnaires administered to patients that assess their physical functioning), performance outcome measures^6^ involving measurement observation of a patient’s function (e.g. 6-minute walk test [6MWT], Timed Up and Go [TUG] test), and physiologic and functional data collected using digital health technologies such as wearable sensors. Given that there are multiple approaches to assessing PF, quantitative data are needed to understand differences in measurement characteristics between these distinct data sources, including variability over time, agreement among measures, sensitivity to changes, and meaningful levels of change.

### Historical approach to evaluating physical function in cancer clinical trials: clinician-reported assessment (ClinRo)

The widely accepted method for recording a patient’s overall functional status in most cancer clinical trials has historically been clinician-or investigator-reported PS using scales such as the Karnofsky performance status (KPS)^7^ and its derivative, the ECOG PS^5^. These tools have become a ubiquitous, international standard in hematology/oncology practice and research. While the simplicity of the PS is attractive, it is also a drawback, as it lacks granularity, which becomes particularly relevant in the setting of patients at ECOG PS 2-3 and clinical trial eligibility. Many trial eligibility criteria exclude patients with ECOG PS ≥ 2, thus leaving the subjective judgement of an oncologist as the main factor determinant of whether a patient can receive what is often a highly desirable therapy on study, or not^8^. This lack of granularity may also impact its sensitivity as a longitudinal outcome measure of changes in PF, reducing the utility of this measure, originally developed as a prognostic tool, when used as a clinical trial outcome assessed over time. Additional limitations to the ECOG PS as a longitudinal measure of physical functioning are that the score is clinician-assessed, rather than directly reflecting the patient experience^9^, and is rarely assessed post-baseline in most cancer trials.

### Novel and more comprehensive approaches to measuring PF which complement ClinRo

#### Patient-reported outcomes (PROs)

While patient health records and provider assessments are invaluable resources for clinical care and research, the patient’s voice is most often absent. PROs are reports of the status of the patient’s health that come directly from the patient, without interpretation of the patient’s response by a clinician or caregiver^10^. PROs are an assessment method that can be used to directly capture many aspects of a patients’ health, from individual symptoms to functional domains such as physical-, emotional-, cognitive, and social function, to the broad multi-domain concept of health-related quality of life (HRQOL). Only patients can tell us how their treatments affect their well-being, as every patient has different goals, values, and preferences. Despite advances in cancer care and delivery, many patients with cancer experience substantial symptoms from disease, side effects from treatment, and functional decline that negatively affect their HRQOL. Clinicians often miss or underreport symptomatic adverse events (AE) experienced by patients that can lead to physical, psychological, and other toxicities going unrecognized^11 12^. The systematic incorporation of PRO assessment to measure symptoms and function that affect patients’ HRQOL in cancer clinical trials is now recognized as critical to complement standard tumor, survival, and clinician-reported safety data by patients, clinicians, industry, academics, and regulators. ^13-15^

Some of the more commonly used PRO measurement systems used in cancer research include the European Organisation for Research and Treatment of Cancer (EORTC) questionnaires^16^, Patient-Reported Outcomes Measurement Information System (PROMIS) questionnaires^17^, and the Functional Assessment of Chronic Illness Therapy (FACIT) questionnaires^18^. Several of these tools include items or subscales that assess physical functioning. The Patient-Reported Outcomes version of the Common Terminology Criteria for Adverse Events (PRO-CTCAE)^19^ is a library of important symptomatic adverse events that can quantify symptomatic toxicities from the patient perspective and can inform causative symptoms that may impact physical functioning. Additionally, prior studies have demonstrated the benefit of patients (in addition to clinicians) directly reporting their own ECOG PS^20^, and patient-friendly versions of the ECOG PS are available^21-23^.

The Patient Global Impression scales (“of change” abbreviated as PGI-C; or “of severity” abbreviated as PGI-S) are single item questions used to evaluate the patient’s perception of change in PF and severity.^24^ These questions are often used to assess meaningful change in PRO scores and other functional measures. There are also questions that are disease-specific, and tools designed to focus more specifically on a particular domain such as physical function.^25^

#### Performance outcome (PerfO) measures

PerfO measures are defined as a measurement based on standardized task(s) actively undertaken by a patient according to a set of instructions. A <u>PerfO</u> assessment may be administered by an appropriately trained individual or completed by the patient independently.^6^ There are a variety of validated PerfO measures that can be used to more objectively measure a patient’s physical PS, including the TUG test, the Sit-Rise test, the Short Physical Performance Battery, gait speed, and grip strength.^26 27^ The TUG has been used to predict falls in a cohort of geriatric patients with cancer, but the others have not been validated in broader cancer cohorts.^28^ As these tools are primarily used in geriatric populations, they may not be as discriminating with younger patients who have better baseline physical fitness.

On the other hand, the 6MWT is a comprehensive measure of exercise capacity suitable for a broad age range. The 6MWT encompasses components of mobility, endurance, and functional capacity.^29-31^ It is relatively straightforward to administer, requires little expertise or training for the patient, and involves minimal equipment. The 6MWT has been used in patients undergoing cancer treatment as well as cancer survivors^32 33^ and normative values for patients with hematologic malignancies have been published. In this study, the standard, validated 6MWT has been selected as the PerfO of interest.

#### Wearable technologies

Wearable products have steadily advanced over the last several years with rapidly evolving sensor technology to measure human movement, such as accelerometers, magnetometers, and gyroscopes.^34^ Commercially available, consumer-grade wearables capable of tracking movement have become ubiquitous to the general public in recent years.^35^ These products can further inform our measurement and understanding of PF by allowing passive monitoring of physical activity in the real world setting. Wearable technology mitigates some of the limitations of self-reported data (e.g., avoiding recall bias), and the narrow validity of data generated in tightly controlled research lab environments.^34 36^

Wearables have been used to assess physical rehabilitation of patients with disabilities and elderly or hospitalized patients.^37-40^ Both capacity (what a patient can do, such as maximal gait speed) and performance (what a patient does, such as total steps per day) have been measured using wearables when assessing changes in PF.^35^ A recent study demonstrated a correlation of heart rate variability measured through a wearable product with PF assessed using the Short Physical Performance Battery scores, TUG scores, and self-reported PF (SF-36 physical composite scores).^41^ The correlation of average daily steps with the 6MWT, another established capacity assessment, was also reported by a recent study.^42^

Fitbit activity tracking products were selected for this study as they have demonstrated acceptable accuracy for heart rate, step count and moderate to vigorous physical activities (MVPA) when compared to research-grade tracking products.^43-45^ Additionally, they are widely available and familiar to consumers.

### Unmet needs in the evaluation of physical function in cancer patients

There is an unmet need to better characterize the measurement characteristics of ClinRo, PRO, PerfO and wearable data to inform selection of measures to meet individual cancer clinical trial objectives. For most therapeutic trials, it may be sufficient to select a single measure suitable across a wide variety of trial contexts to foster standardization, while comparative tolerability trials may use several measures to increase confidence in findings. In all cases, a firm scientific understanding of measurement characteristics including variability, sensitivity, and meaningful change across all modalities would advance our ability to make science-driven trial design decisions and best inform regulatory and clinical decision-making. Operational aspects including ease of use and adherence are also critical to identify methods to reduce missing data-a key challenge to interpreting PF results regardless of assessment modality.

Few studies have demonstrated the logistical feasibility, sensitivity, and complementarity of different PF measurement modalities in the cancer treatment context. There has been no clear identification of meaningful levels of change for these measures either with respect to patient experience or in correlation with adverse event or hospitalization rates. Such data would inform potential use of PROs and digital hardware in the design of tolerability endpoints for regulatory review in cancer clinical trials in all phases of medical product (i.e., drug, device, and biologics) development.

In this prospective study, we will evaluate PF in patients with cancer undergoing routine treatment. We will collect PF data across four assessment modalities in a population of patients with solid tumors and hematologic malignancies receiving cytotoxic chemotherapy with standard clinical follow up and care.

#### Study Aims

The purpose of this study is to integrate four PF measures (ClinRo, PRO, PerfO and wearable data) in a prospective cohort of patients receiving chemotherapy for breast cancer or lymphoma. Using a digital health-based patient-centered data aggregation platform, Hugo Health, we aim to collect and compare PF trajectories and establish measurement characteristics for the different assessment modalities of PF.

There are three main study aims:

1. To measure PF using ClinRo, PRO, PerfO and wearable data. This includes characterizing feasibility and assessment challenges by comparing levels of missing data and reasons for missingness across the PF modalities and report on trajectories of function as ascertained by the four PF modalities.
2. To explore associations between various sources of PF data and determine meaningful change thresholds. This includes assessing measurement characteristics of the different modalities, including sensitivity to change and identification of meaningful change thresholds; comparing changes within and between modalities; and exploring associations between changes in the PF modalities and subsequent clinical outcomes, such as patient-reported AEs, other patient-reported domains of HRQOL, acute care usage, and chemotherapy dose delay/reduction.
3. To assess patient acceptability and experience using the different PF assessment modalities, via the use of an exit questionnaire, to understand burden and usability of electronic PROs and wearable data collection from the patient perspective.

## METHODS

In this prospective study, we are collecting PF data across the four different assessment modalities in a population of patients with breast cancer or lymphoma receiving routine-anticancer therapy including a cytotoxic chemotherapy. We plan to follow patients prospectively for 9 months, tracking clinician and patient self-report of physical functioning, PerfOs, and wearable data using a patient-centered health data sharing platform – Hugo Health – that will consolidate data from electronic health records (EHR), patient surveys, and wearable data (See Figure 1, Study Schema). Patients use their personal smartphone or other web-connected mobile product to answer questionnaires about PF, symptoms and adverse effects. Information from the EHR is collected to record baseline clinical features, clinician-reported performance status, treatment plans, and outcomes including acute care usage (emergency department visits, hospitalizations), and chemotherapy dose reductions, delays or discontinuations.

**Figure 1.**
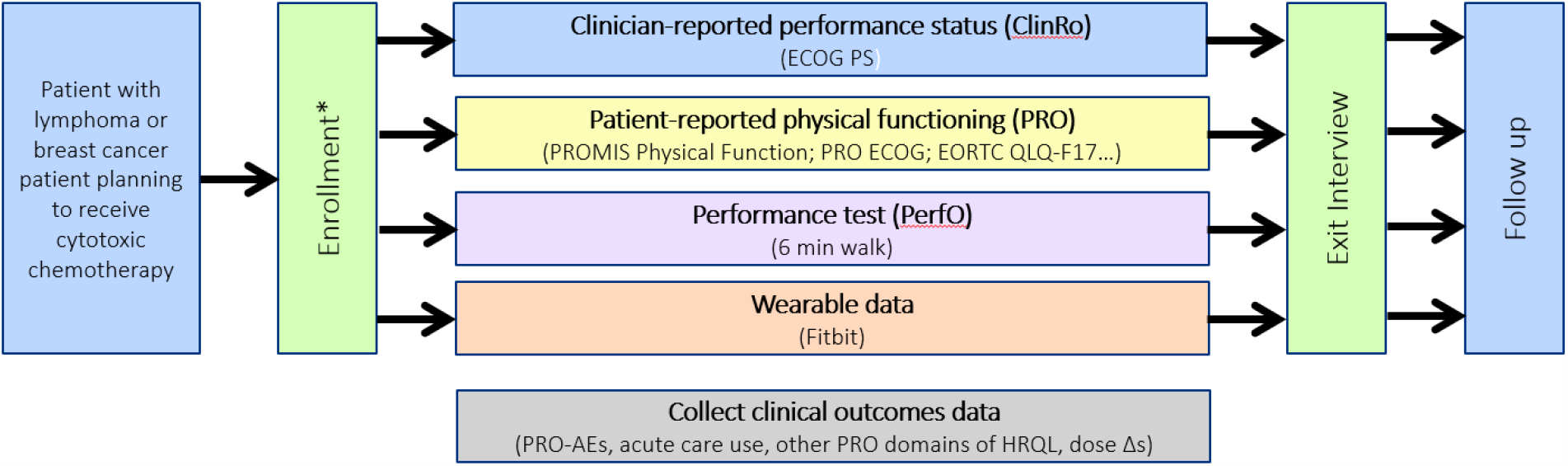
Study Schema

The study is based at Mayo Clinic (Minnesota) and Yale University. Participants are recruited both at community and academic hospitals, as well as clinics affiliated with these sites. Participants can be treated after recruitment at a local community site and followed remotely after study consent and enrollment is obtained at the primary site. Informational flyers are placed in waiting rooms of breast cancer and lymphoma clinic practices at both primary sites.

Charts of potential study candidates are reviewed by clinical investigators, and if potentially eligible, patients are approached about and consented for the study by the study research assistants. Each site will enroll 100 patients. Complete inclusion and exclusion criteria are in Appendix 1.

### Measures and Data Collection

A detailed description of Hugo Health, the electronic health data aggregating technology used to administer PRO questionnaires, collect patient electronic health record portal data, and aggregate wearable data in this study, has been published previously.^46 47^ All of the data and records described below and generated during this study are kept confidential in accordance with institutional policies and the Health Insurance Portability and Accountability Act (HIPAA) on subject privacy.

### Clinician-Reported Performance Status (ClinRo) and Performance Outcomes (PerfO)

Clinician-reported performance status is recorded from the medical record into a REDcap form by research assistants every 3 months. The 6MWT is performed once at baseline (prior to start of chemotherapy) and at 3 months for participants treated at Mayo Clinic and Yale primary sites. Participants receiving care at another site will not have an additional 6MWT observation.

### Patient-Reported Outcomes (PRO)

Questionnaires are sent by Hugo to patients throughout the 9-month follow-up period (Table 1). To inform our measurement approach, we engaged three patient advocate co-investigators who reviewed the schedule of assessments to minimize participant burden. PROs assessing PF include the PROMIS version 2.0 physical function 8c short form, PF questions from the EORTC QLQ-F17 instrument, a patient-adapted version of the ECOG PS (PRO-ECOG), and the PGI-C/PGI-S items pertaining to PF. Additional PROs that capture global assessments of quality of life and well-being (functional and QOL domains of the EORTC QLQ-F17 and selected items from the PRO-CTCAE, FACIT GP5) are used to assess the correlation of PF data with symptomatic toxicities, patient-reported AEs, and other domains of HRQOL. Hugo sends automated reminders if patients do not complete the weekly survey after 48 hours or the monthly survey after one week. Additionally, at key timepoints, research assistants call patients if questionnaires have not been completed after 5 days for weekly questionnaires or after 2 weeks and 2 days for the monthly questionnaires.

**Table 1.**
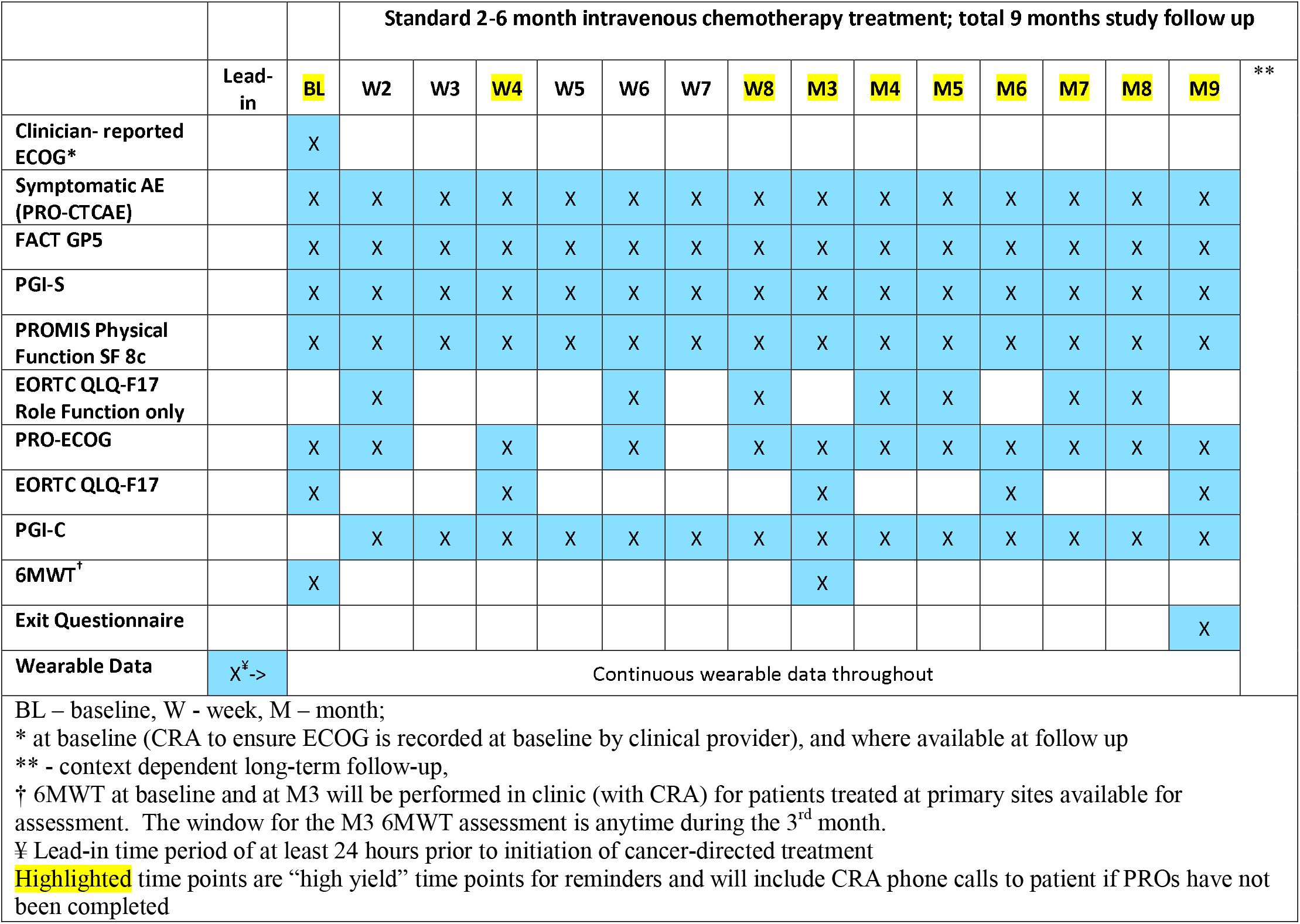
Schedule of Assessments

### Wearable data

A Fitbit model with built-in GPS, the Fitbit Inspire, is used in this study. Multiple data parameters are recorded from the lead-in time point to the completion of month 9 of follow up. The lead-in time, for baseline data collection prior to initiation of cancer-directed therapy, was pragmatically derived to be at least 24 hours. Fitbit data are automatically uploaded from the wearable to Fitbit’s servers when the Bluetooth feature on the patient’s wearable is turned on. Hugo downloads that data through the Fitbit API regularly and links it to the other participant data. All wearable data is collected and stored via Hugo.

Patients are instructed to (1) wear the Fitbit as much as possible during the day and night, limiting non-wear time to recharging periods (approximately 1-2 hours every 3 days) and (2) synchronize (upload) the Fitbit data from the wearable to Fitbit’s servers every 3 days using the Fitbit smartphone application. Reminders to synchronize Fitbit data are delivered by Hugo to study participants on a weekly basis. Additionally, Fitbit data are reviewed for completeness by the study team weekly and patients whose data has not been received are contacted by research assistants.

Predefined parameters evaluating both capacity and performance measurements of PF from three domains (steps/distance, heart rate, and activity level) will be used for comparison with the other PF assessment modalities. Additional metrics of interest derived from the raw data parameters or obtained directly from Fitbit will be considered. These additional metrics may include distance walked per day, sleep duration per day, heart rate variability, sleep cycle duration, etc.

### Analysis Plan

Specific Aim 1: In order to characterize assessment challenges, completion rates will be computed and reasons for missing data will be described. For each PF metric, the completion rate will be computed at applicable time points using (1) a fixed denominator method using all patients ever enrolled, and (2) a variable denominator method using the number of active patients at each time point. For the variable denominator approach, at each timepoint, active participants are those who have not died and have not withdrawn from study participation. Intercurrent events including reason for study withdrawal, disease progression, and death will be summarized in analysis.

To describe distributions of PF responses over time, the trajectory of each PF metric will be graphically explored using stream (spaghetti) plots and mean plots. Mean plots will employ raw means as well as estimated means from a general linear mixed modeling at each time point. Estimation will include group means and group mean changes from baseline.

Specific Aim 2: To identify measurement characteristics of each PF metric, standard psychometric analyses investigating sensitivity to change, and meaningful change thresholds will be carried out. These analyses will employ both anchor-based and distribution-based methods. The primary anchor will be PGI-C and the key secondary anchor will be PGI-S.

Distribution-based analyses for each PF metric will include the mean, standard deviation, median, first quartile, third quartile, minimum, and maximum. Effect sizes representing small, moderate, and large effects will be computed as 0.2, 0.5, and 0.8 times the baseline standard deviation.

Anchor-based analyses will estimate the mean change for each PF metric over time according to how patients respond to the PGI-C and PGI-S items. Mean change at each post-baseline timepoint will be described using the mean and standard deviation within strata of patients grouped by their status change (those reporting worsening status; no change in status; and improved status) and their current limitations in PF (no limitations, mild or moderate limitations, and severe limitations). Additionally, the standardized response mean (SRM) will be computed as the mean change score divided by the standard deviation of the change scores within each change category (worsening vs. no change vs. improvement) or severity category (normal vs. mild/moderate vs. severe). Values greater than 0.8 will be considered large and values between 0.5 and 0.8 will be considered moderate. Additionally, Spearman correlations between the change in each PF metric and the change in other anchors (e.g., physician-reported and patient-reported ECOG PS, patient-reported role function, global health status/QOL, and HRQOL via the EORTC QLQ-C17; PRO-CTCAE symptomatic adverse event grades; and FACIT GP5) will be computed.

The relationship between change in PF metrics and PGI-C and PGI-S items will be investigated using general linear mixed models. Mean change from baseline with 95% confidence intervals will be computed for each PF metric based on mixed modeling. Mixed models will include all PF metrics as outcomes and time as a categorical variable. Additional patient or design characteristics will be incorporated as baseline covariates. Composite covariance will initially be used, with the final covariance structure selected based on minimization of the Akaike information criterion. All patients who consent for participation in this study and complete at least two PF metrics will be included in statistical analysis. In the primary analysis, all observations available will be used.

We will conduct secondary analyses, assessing the association between baseline patient characteristics and baseline PF metrics using Spearman correlations and longitudinal PF metrics using statistical modeling. Key baseline patient characteristics that will be explored as feasible based on the distribution of the characteristics observed in the sample will include. cancer cohort (breast vs. lymphoma); age (<65 vs. ≥65 years); physician-reported ECOG PS; patient-reported ECOG PS; patient-reported role function, global health status/QOL, and HRQOL via the EORTC QLQ-C17; PRO-CTCAE symptomatic adverse event grades; and FACIT GP5.

Association between longitudinal patient characteristics (patient-reported ECOG PS; patient-reported role function, global health status/QOL, and HRQOL via the EORTC QLQ-C17; PRO-CTCAE symptomatic adverse event grades; and FACIT GP5) and longitudinal PF metrics will be explored using Spearman correlations at successive time points as well as statistical modeling (bivariate linear mixed modeling).

Specific Aim 3: Statistical analysis will be primarily descriptive for the exit questionnaire data. Continuous outcomes will be summarized using means, standard deviations, medians, minimums, and maximums. Categorical outcomes will be summarized using frequencies and relative frequencies.

Power considerations: Our targeted sample enrollment is 200 patients, which we expect will allow the team to have data available for a given PF metric at early post-baseline timepoints (at least the first 3 months) for at least 170 patients. Based on a prior study evaluating association between PF as measured by the QLQ-C17 and a PGI-C item assessing physical condition^19^, we anticipate 25% of patients to report worsening and the mean change in PF among these patients to be -8.2 points. The remaining 75% of patients reporting no change or improvement had a mean change in PF of 0.9 points (pooled standard deviation 15.0). Thus, with a sample size of 170 patients, this study has 92% power to detect a similar change as the prior study using a t-test comparison with a two-sided alpha of 0.05. Statistical analysis will employ a modeling approach across all time points and thus power estimation based on a single time point can be considered conservative.

## Missing data

Missing data from patient questionnaires will be handled in a number of ways. Missing items within a summary or scale score will be handled according to each questionnaire’s published scoring algorithms. When summary or scale score data are missing, baseline patient/disease characteristics will be compared between patients who do and do not provide data for a given analysis and patterns of missing data will be graphically explored. All analyses will first be completed using all available data, then by integrating missing categories for categorical data and analyses completed using multiple imputation via chained equations (20 or more for each analysis), and finally using pattern mixture models for longitudinal analyses. Output from all analyses will be tabulated and descriptively compared to assess the degree to which missing data impacts study results.

For all statistical analyses, p-values <0.05 will be considered statistically significant; however, interpretation will take into consideration that type I error is not strictly controlled across all planned analyses. For interpreting the clinical significance of effects, 0.2, 0.5, and 0.8 standard deviation (SD) effects will be considered as small, moderate, and large.

### Data collection and management

The Hugo platform will aggregate data from the EHR, PROs and wearables. At study enrollment, patients provide Hugo access to their health portals by authenticating themselves using their username and password. PerfO and clinician-reported ECOG will be among data collected by the research assistant and entered into a secure REDCap database. Additionally, clinical co-investigators will review the medical records of each patient directly for more granular information on tolerability parameters, such as reasons for hospitalizations or dose reductions, and these data are entered into the study REDCap database by the research assistant.

### Patient involvement

Three patient-advocate co-investigators provided input on the design of the study, the selection of PRO survey items, and timing of scheduled assessments. They also co-created a “study welcome letter” to describe in patient-tailored language the purpose of the study, and they have participated in the writing and review of this manuscript. Patient advocates were not involved in the conduct of the study.

### Study limitations

Although patients on this study can receive their cancer treatment at primary or local sites as part of this clinical study, recruitment is limited to patients seen at least once at Mayo Clinic or Yale clinical sites, limiting participation to patients who have the physical and financial ability to access these tertiary cancer care centers. Most participants receive treatment at the primary sites and may not be representative of a larger community oncology practice. We do not offer patients a smartphone or other web-connected product if they do not have one, which may limit participation, though smartphone adoption is high at 85% of American adults, including a majority of those with low income and those living in rural areas, with minimal gaps by race and ethnicity.^48^ Some patients who already use a non-Fitbit wearable product or are apprehensive of wearable data collection may decline participation. Lastly, we do not have formalized technology support for patients over and above the research assistants in this study, which may limit our ability to swiftly address technical issues related to Hugo or Fitbit.

### Ethics and Dissemination

Institutional review board (IRB) approval was secured at Mayo Clinic, Yale University and the U.S. Food & Drug Administration. Study results will be disseminated through publications in general, and specialty medical journals and conferences.

### Study Update

At the time of this publication, all sites have obtained local IRB approval and are enrolling participants. The COVID-19 pandemic delayed study activation at both sites; enrollment in this study began in January 2022. 123 participants have been enrolled at the time of this manuscript submission.

## Data Availability

N/A

## Authors’ contributions

Conception or design of the work: PGK, GT, CPG, VB, MMJ, MD, MT, JSR, JDR, AB, LJ, BP, ACD, KJR.

Planning for acquisition, analysis or interpretation of data: MD, LF, BNN, JDR, MF, ACD. Screening, enrollment and health record review: GT, ECR, SH, JP, KJR, SES, CPG. Drafting the work: GT, CPG.

Revising the work critically for important intellectual content: All authors

Final approval of the version to be published: All authors.

Agreement to be accountable for all aspects of the work in ensuring that questions related to the accuracy or integrity of any part of the work are appropriately investigated and resolved: GT and CPG.

## Competing interests

Dr. Thanarajasingam has received research funding from the from the Food and Drug Administration for the Yale-Mayo Clinic Center of Excellence in Regulatory Science and Innovation (CERSI) (U01FD005938) that directly supports this work. She also receives grant funding from the National Cancer Institute (NCI) U01 Tolerability Consortium.

Dr. Gross has received research funding from the NCCN Foundation (Astra-Zeneca) and Genentech, as well as funding from Johnson and Johnson to help devise and implement new approaches to sharing clinical trial data.

Over the past three years, Dr. Jeffery reports grant funding from the US Food and Drug Administration, National Institutes on Drug Abuse, Centers for Disease Control and Prevention,

Agency for Healthcare Research and Quality, American Cancer Society, and the National Center for Advancing Translational Sciences.

Dr. Ross currently receives research support through Yale University from Johnson and Johnson to develop methods of clinical trial data sharing, from the Food and Drug Administration for the Yale-Mayo Clinic Center of Excellence in Regulatory Science and Innovation (CERSI) (U01FD005938), from the Medical Devices Innovation Consortium as part of the National Evaluation System for Health Technology (NEST), from the Agency for Healthcare Research and Quality (R01HS022882), from the National Heart, Lung and Blood Institute of the National Institutes of Health (NIH) (R01HS025164, R01HL144644), and from Arnold Ventures for the Collaboration for Regulatory Rigor, Integrity, and Transparency (CRRIT); in addition, Dr. Ross is an expert witness at the request of Relator’s attorneys, the Greene Law Firm, in a qui tam suit alleging violations of the False Claims Act and Anti-Kickback Statute against Biogen Inc.

Ms. Ritchie currently receives research support through Yale University from Johnson & Johnson to develop methods of clinical trial data sharing and from the US Food and Drug Administration for the Yale-Mayo Clinic Center of Excellence in Regulatory Science and Innovation (CERSI) (U01FD005938).

Dr. Huntington has received consulting fees outside of this work from Janssen, Genentech, AbbVie, Flatiron Health, BeiGene, AstraZeneca, ADC Therapeutics, Epizyme, Merck, Seattle Genetics, TG Therapeutics, Tyme, Pharmacyclics, SeaGen, and Arvinas.

Dr. Schellhorn has received consulting fees from Eisai, Celgene, SeaGen, and Cardinal Health. She has previously received research funding to her institution from Genentech and Pfizer.

All other authors have no relevant conflicts of interest to disclose.

## Appendices

Appendix (Supplementary Material) 1: Inclusion and Exclusion Criteria for the In4M Study

### 1.1.1. Inclusion Criteria

1. Age 18 and over;
2. English-or Spanish-speaking;
3. Pregnant and non-pregnant patients are eligible for participation in this study
4. Eligible cancer type and planned intravenous cytotoxic chemotherapy regimen (defined as including 1 or more cytotoxic agents)
5. ECOG Performance Score of < 3
6. <u>Breast cancer patients</u>
  a. Patients with any stage breast cancer for whom a new intravenous cytotoxic chemotherapy regimen is planned within the next 8 weeks (patients with local/regional/distant recurrences are allowed; patients with concurrent/prior/future immunotherapy/radiotherapy, targeted therapy, and endocrine therapy for breast cancer are allowed)
7. <u>Lymphoma patients</u>
  a. Lymphoma patients of any histology, stage or line of treatment planned to receive a new intravenous cytotoxic containing chemotherapy regimen (patients planned to receive radiation, maintenance chemotherapy, consolidation stem cell transplant or chimeric antigen receptor T (CAR-T) cell therapy are allowed)
8. If patients are receiving the above standard therapies as part of a clinical trial which may include a novel agent or combination, they are also eligible for the present study if the therapeutic protocol permits enrollment in both studies
9. Willing and able to give consent and participate in study
10. Able to access a mobile smartphone or tablet or computer with web access every day to complete study surveys; able to regularly upload data from the Fitbit to a in a way that it can be transferred to Hugo.
11. Willing and able to perform an in-clinic 6-minute walk test (gait aides are permitted if regularly used by the patient). If a patient is recruited remotely outside of Mayo Clinic Rochester or Yale Smilow Cancer Center New Haven, 6-minute walk test may be omitted.
12. Willing to use the health data sharing platform Potential subjects who do not meet all of the enrollment criteria will not be enrolled. Any deviations from these criteria must be reported in accordance with IRB Policies and Procedures.

### 1.1.2. Exclusion Criteria

1. Prior intravenous cytotoxic chemotherapy within 3 weeks prior to study enrollment
2. Excluded regimens (due to length of hospitalization required for chemotherapy administration):
  a. R-CODOX-M/IVAC,
  b. DA-R-EPOCH (inpatient)
3. Excluded histology (due to length of hospitalization and inpatient predominant treatment for required chemotherapy): primary central nervous system lymphoma
  a. Other regimens with an anticipated high duration of inpatient care time, at PI discretion
4. Lack of access to a mobile smartphone or tablet or computer with web access
5. Unable or unwilling to upload data from the Fitbit
6. Unable or unwilling to use the health data sharing platform
7. Unable to give consent and be enrolled

